# Mood instability may be causally associated with the high risk of cardiovascular disease: evidence from a mendelian randomization analysis

**DOI:** 10.1101/2023.08.29.23294761

**Authors:** Zirui Liu, Haocheng Wang, Zhengkai Yang, Yu Lu, Jikai Wang, Cao Zou

## Abstract

**Background:** Mental illness was identified associated with high risk of cardiovascular diseases (CVDs). However, few studies focused on the effect of personality traits, the causal relationships remain unknown. Here, we use mendelian randomization (MR) analyses to evaluate the causal association between mood instability (mood swings) and 5 common CVDs.

**Methods:** Large genome-wide association studies (GWAS) of mood swings (n= 373733) and 5 CVDs from two independent cohorts respectively including coronary artery disease (CAD) (n= 766053), myocardial infarction (MI) (n= 596436), heart failure (HF) (n= 1185501), atrial fibrillation (AF) (n= 2169833) and stroke (n = 627558). We performed a range of bidirectional two-sample MR and related sensitive analysis including MR-Egger regression, MR-PRESSO global test and “Leave-one-out” method. A Bonferroni-corrected significance level of p < 0.01 (0.05/5) was identified to be statistically significant, while p < 0.05 was considered to indicate suggestive evidence. Moreover, multivariable MR (MVMR) and mediation analyses were also conducted to adjust confounding factors as well as found potential mediators.

**Results:** This MR analyses revealed the significant causal effects of mood swings on CAD (OR = 1.45, 95% CI 1.24–1.71; P = 5.52e-6), MI (OR = 1.60, 95% CI 1.32–1.95; P = 1.77e-6), HF (OR = 1.42, 95% CI 1.12–1.71; P = 2.32e-6) and stroke (OR = 1.48, 95% CI 1.19–1.83; P = 3.46e-4). However, no causal effects of mood swings on AF (P=0.16) were found. In the reverse MR, no causal relationships were observed. Additionally, hypertension may mediate the causal pathway from mood swings to CAD (proportion of mediation effect in total effect: 39.60%, 95% CI: 19.31%–59.89%), MI (35.37%, 95% CI: 17.10%–53.65%), HF (43.19%, 95% CI: 20.68%–65.69%) and stroke (55.47%, 95% CI: 27.00%–83.95%).

**Conclusion:** Mood instability (mood swings) causally resulted in CAD, MI, HF and stroke, and these causal effects may be partly mediated by hypertension.

## 1. Introduction

Mood instability, a personality trait presents in normal individuals, is defined as a frequent, sudden and unpredictable alteration in emotional states^1^. Mood instability is relatively common in the general crowd—approximately 45% participants in the UK biobank (n=488105) admitted that they experience frequent fluctuations in their emotional state^2^, such as anger transitions into sadness or anxiety, excitement to disinterest, or happiness turns into tearful sorrow. Although mood instability is not a psychotic disorder, many studies have identified its tight association with mental illness including major depressive disorder (MDD), bipolar disorder (BD), schizophrenia and anxiety disorder^3-5^.

Numerous research has focused on the relationship between psychotic illness and common cardiovascular diseases (CVD) such as coronary artery disease (CAD), atrial fibrillation (AF), heart failure (HF), myocardial infarction (MI) and stroke. Similar conclusions were drawn from different studies that psychotic illness could not only increase the risk of CVD, but also be associated with high risk of all-cause and cardiovascular disease mortality^6-9^. However, limited studies focused on the relationship between personality traits (such as mood instability) and CVD.

In contrast to psychotic disorders that are usually late onset, after birth personality traits start to exert a sustaining influence on people’s behaviors and biological processes^10^, which may play an essential role in the progress of CVD. So, we hypothesize that mood instability as an upstream factor for psychotic disorders may exert a profound impact on the progress of CVD. Yaa A. Kwapong^11^ et al. used “poor mental health days (PMHD)” to assessment the potential relationship between PMH and CVD. Although they found that more PMHD brought more risk of CVD to participants, this cross-sectional study cannot provide any causal relationship or the directionality of the association.

Mendelian randomization (MR) is a method employing genetic variants as instrumental variables (IVs) to verify whether exiting a causal relationship between exposures and outcomes^12^, which based on the principle of gene random segregation and independent assortment during the process of meiosis^13^. Because MR is not affected by environmental factors, it effectively avoids confounding factors and reverse causality, which are always inevitable in observation studies^14^. Recent MR studies have focused on the causal effect from mental illness to CVD^9,15,16^, fully clarifying the viewpoint that the risk of CVD is significantly higher than those without psychotic disorder. However, whether mood instability may causally associate with the high risk of CVD is still unknown.

In this article, we preformed the first bidirectional two-sample MR analysis to investigate the causal association between mood instability and 5 common CVDs, including CAD, MI, HF, AF, and stroke by using genome-wide association studies (GWAS). Two independent cohorts were estimated and pooled to ensure the robustness of the casual effects. Additionally, we further explored whether the casual relationships of mood instability on CVDs is mediated by hypertension by using mediation MR analysis.

## 2. Materials and methods

### 2.1 Study design

In this study, we use the bidirectional two-sample MR method to clarify the casual relationships between mood instability and 5 common CVDs with GWAS summary statistics. In addition, we conducted multivariable MR (MVMR) and mediation MR analysis to eliminating the influence of confounding factors and investigate whether any regular risk factors of CVD, such as smoking, drinking, body mass index (BMI), Interleukin-6 (IL-6), C-reactive protein (CRP) and hypertension may mediate the casual effect. The framework of this study was shown in **Figure 1**.

**Figure 1.**
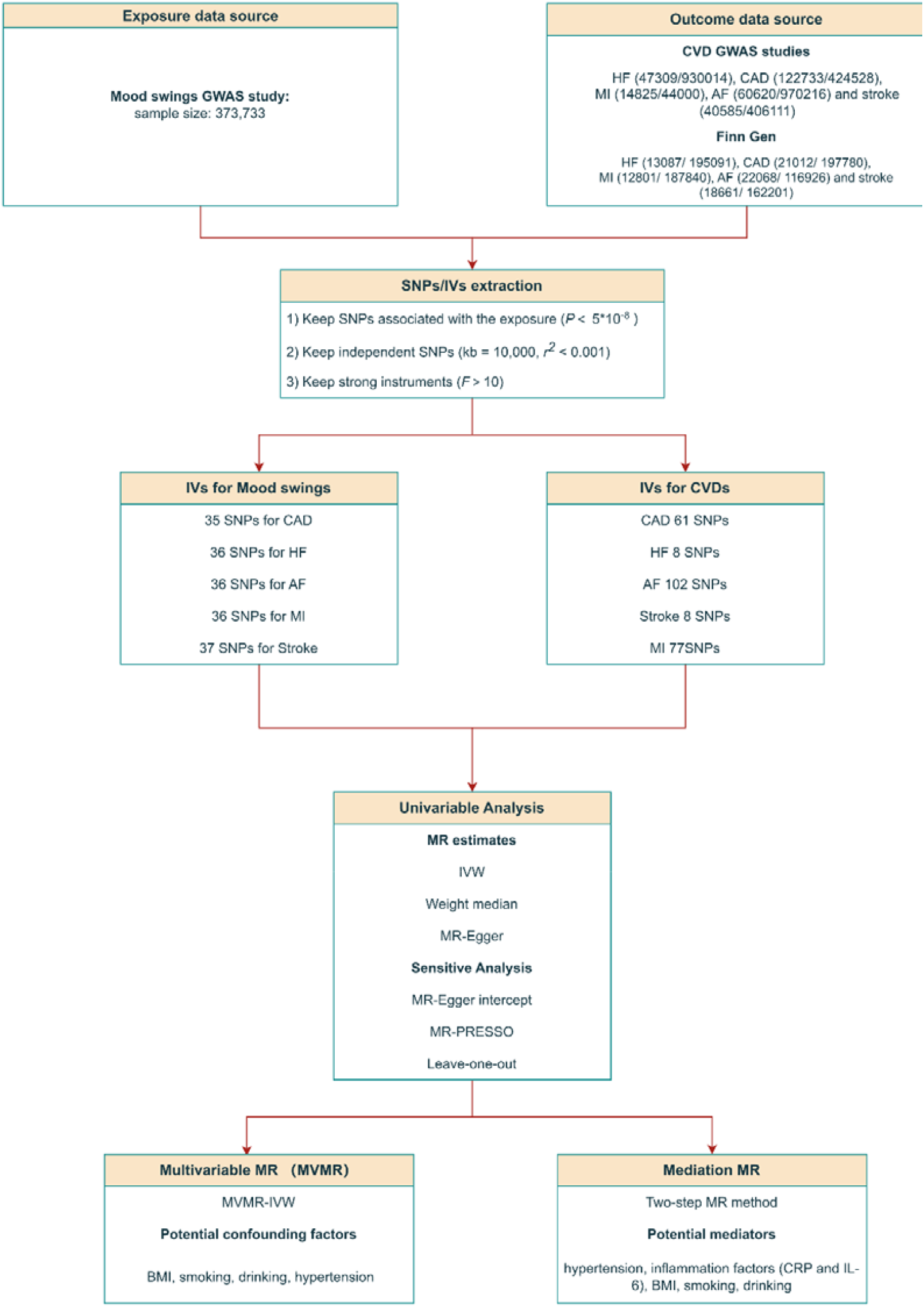
Study design of MR study

All statistics were calculated using R software 4.2.2 (The R Foundation for Statistical Computing).

### 2.2 Genetic association datasets for bidirectional MR analysis

All the single nucleotide polymorphisms (SNPs) used in this MR study as IVs must satisfy the following 3 core assumptions: 1) genetic IVs are robustly associated with exposures; 2) no confounders are associated with IVs; 3) IVs affect outcomes only by their effects on exposures rather than other pathways. **(Figure S1)**. All the summary statistics were publicly available which obtained from the Open GWAS database developed by the MRC Integrative Epidemiology Unit (IEU) (https://gwasmrcieu.ac.uk/). As all individual studies that contributed to this MR study had obtained the necessary consent and ethics approvals, no additional consent or ethics approval was specifically required.

We selected valid SNPs for this study based on the required MR. Firstly, all SNPs robustly associated with exposures (p < 5e-8) were extracted as candidate IVs. Secondly, we used linkage disequilibrium analysis to obtain independent SNPs (kb = 10,000, r2 < 0.001). Thirdly, the F-statistic was calculated for each SNP to avoid “weak instrument” bias (*F* > 10), the formula is:

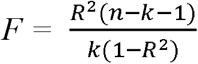

where, n, k, R^2^ represents sample size, the number of SNPs and the variance explained by the instruments respectively^17^. Finally, according to the Bonferroni multiple correction criterion, p < 0.01(0.05/5) was significant, 0.01 < p < 0.05 was considered a potential casual association between exposure and outcome.

#### 2.2.1 Genetic association datasets for mood instability

For mood instability, this study used “mood swings” large-scale GWAS summary statistics, which included 373,733 individuals in UK biobank and focused on 12 neuroticism items^18^.

#### 2.2.2 Genetic association datasets for CVDs

Summary-level data for patients with 5 common CVDs were extracted from the EBI database.

To guarantee the homogeneity of the study population and the reliability of the results, each cardiovascular disease (CVD) was derived from two independent large-scale cohorts. So, we also extracted summary-level data for each CVD from the FinnGen consortium (https://www.finngen.fi/en). All the detailed information were shown in **Table 1**.

**Table 1.**
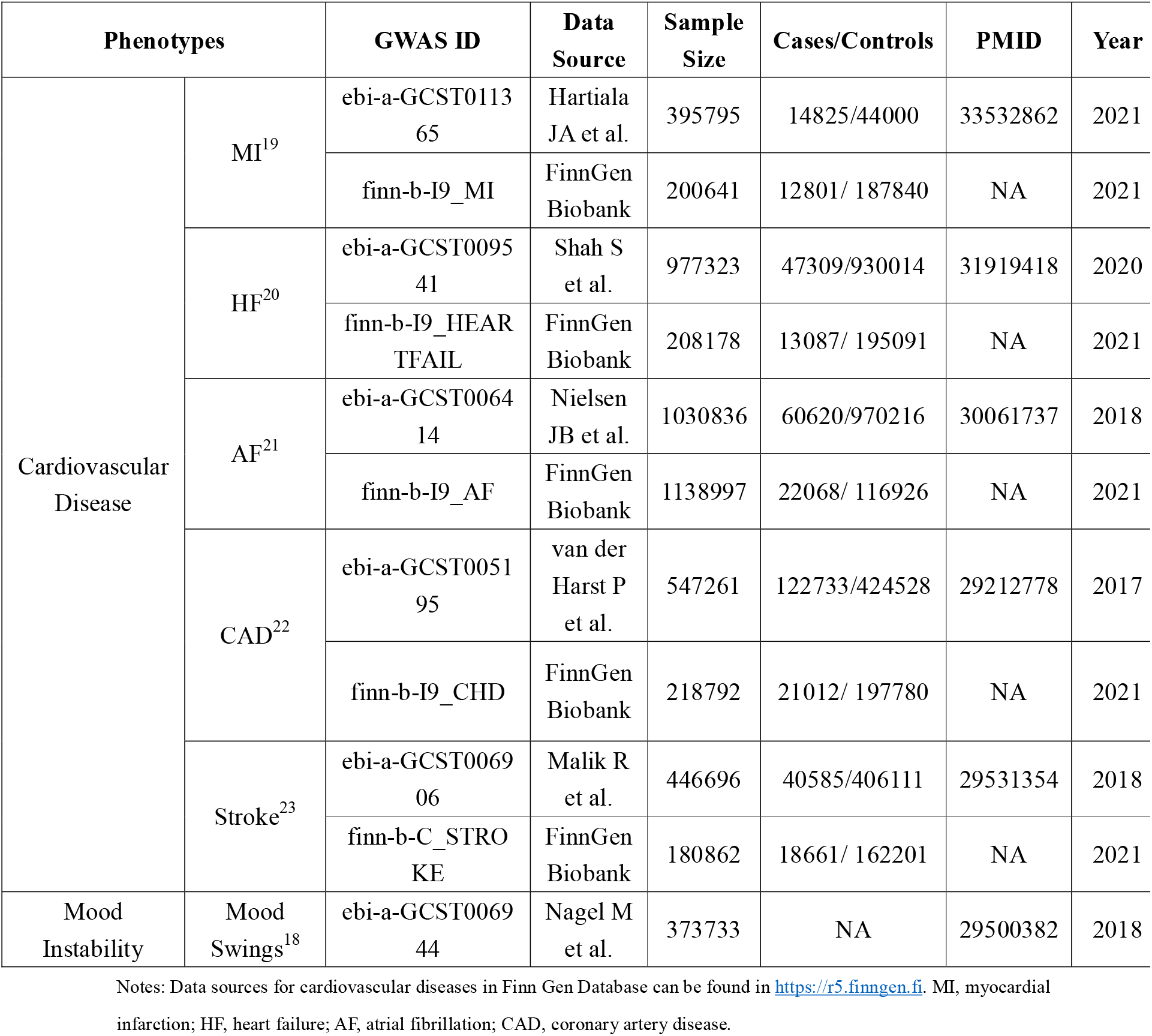
Detailed information from GWAS database.

### 2.3 Statistical analysis

The “TwoSampleMR” package^24^ was used for estimating the casual effects between mood instability and CVDs. The effects were represented by odds ratios (OR) with 95% confidence intervals (CI). Furthermore, we detected and removed outliers by using the “MR-PRESSO” package. The influence of confounding factors was eliminating by using “MendelianRandomization” package^25^, which can also be used for performing mediation analysis.

#### 2.3.1 Univariable MR analysis

In the univariable MR analysis, we used the classical random effect inverse variance weighted method (IVW)^26^ to estimate causal effect between mood instability and different CVD events. Because the horizontal pleiotropy always exists in IVW estimates, we also employed two alternative methods including MR-Egger regression^27^ and Weighted median^28^ for MR analysis to evaluate the robustness of these outcomes. Causal effects were significant if the p value was less than the Bonferroni-corrected threshold (p<0.05/5=0.001) in IVW as well as the direction of these results from the weighted median and MR-Egger were consistent. P < 0.05 but above the threshold was defined as potential causal associations. Then, three complementary methods including MR-Egger intercept, MR-PRESSO (MR Pleiotropy Residual Sum and Outlier)^29^ and Leave-one-out were conducted as sensitive analysis. The MR-PRESSO could recognize potential outlier SNPs (p < 0.05) and correct them before re-estimating the causal effects to examine whether the causal effects would change after removing these outliers. Moreover, the Leave-one-out method was also conducted for evaluating the robustness of the univariable MR analysis outcomes through outlier SNPs. Additionally, for ensuring the homogeneity and reliabilities of these results, we pooled the outcomes from two independent large-scale cohort by means of meta-analysis.

#### 2.3.2 Multivariable MR (MVMR)

We performed MVMR for those significant causal relationships which were identified in the univariable MR analysis to examine whether the outcomes were still significant when eliminating common confounding factors in CVDs (including smoking, drinking, BMI as well as hypertension) by using MVMR-IVW method. If MVMR reveals that any risk factors significantly attenuate the causal effects between mood instability and CVDs (P < 0.005 altered to P > 0.05), it will identify the risk factor as a mediating factor and the mediation effects will be estimated in mediation analysis.

#### 2.3.3 Mediation analysis

Considering that mood instability could cause a range of somatic discomfort symptoms^1^ and previous observational studies have found that hypertension may mediate the pathway to different CVD events^6,11,30,31^, we performed the mediation analysis through the two-step MR method^32^.

Furthermore, any risk factor suggested as a mediation factor in MVMR will also be calculated mediation effects. In the first step, we calculated the causal effect of mood instability on mediators (β1), while in the second step, we estimated the causal effect of mediators on various CVDs (β2). The statistical significance of the mediating effects (β1*β2) as well as the proportion of the mediation effect in relation to the overall effect were estimated employing the delta method.

## 3. Outcomes

### 3.1 Primary analysis

#### 3.1.1 Causal association between mood instability and 5 common CVDs

Firstly, we investigated the causal effect of genetically predicted mood instability on CAD, MI, HF, AF and stroke. The IVW method provided the primary results, which showed that mood instability was strongly associated with CAD (OR = 1.45, 95% CI 1.24–1.71; P = 5.52e-6), MI (OR = 1.60, 95% CI 1.32–1.95; P = 1.77e-6), HF (OR = 1.42, 95% CI 1.12–1.71; P = 2.32e-6) and stroke (OR = 1.48, 95% CI 1.19–1.83; P = 3.46e-4). However, causal effect of mood instability on AF was not significant in the primary analysis. **Table 2** presented the outcomes of these results.

**Table 2.** Outcomes about causal effects of mood instability (mood swings) on different CVDs.

| Outcome | GWAS ID | Method | nsnp | Pval | OR | 95%CI | Meta |
| --- | --- | --- | --- | --- | --- | --- | --- |
| CAD | ebi-a-GCST005195 | Inverse variance weighted | 35 | 5.52E-06 | 1.45 | 1.24-1.71 | 1.49(1.30-1.72)<br>$p=2.88\text{e-}08$ |
|  |  | Weighted median |  | 2.47E-03 | 1.43 | 1.14-1.81 |  |
|  |  | MR Egger |  | 3.21E-01 | 1.60 | 0.64-4.00 |  |
|  | finn-b-I9_CHD | Inverse variance weighted | 40 | 1.14E-03 | 1.63 | 1.21-2.19 |  |
|  |  | Weighted median |  | 5.68E-02 | 1.52 | 0.99-2.34 |  |
|  |  | MR Egger |  | 8.58E-01 | 0.84 | 0.13-5.61 |  |
| AF | ebi-a-GCST006414 | Inverse variance weighted | 36 | 1.64E-01 | 1.12 | 0.96-1.31 | 1.42(0.85-2.36)<br>$p=1.82\text{e-}01$ |
|  |  | Weighted median |  | 5.17E-01 | 1.08 | 0.86-1.35 |  |
|  |  | MR Egger |  | 2.04E-01 | 2.04 | 0.69-5.99 |  |
|  | finn-b-I9_AF | Inverse variance weighted | 37 | 4.19E-04 | 1.89 | 1.33-2.68 |  |
|  |  | Weighted median |  | 3.82E-02 | 1.68 | 1.03-2.76 |  |
|  |  | MR Egger |  | 4.08E-01 | 2.71 | 0.26-28.08 |  |
| Stroke | ebi-a-GCST006906 | Inverse variance weighted | 37 | 3.46E-04 | 1.48 | 1.19-1.83 | 1.47(1.23-1.76)<br>$p=1.86\text{e-}05$ |
|  |  | Weighted median |  | 5.45E-02 | 1.36 | 0.99-1.85 |  |
|  |  | MR Egger |  | 2.62E-01 | 2.18 | 0.57-8.27 |  |
|  | finn-b-C_STROKE | Inverse variance weighted | 37 | 1.87E-02 | 1.46 | 1.07-2.01 |  |
|  |  | Weighted median |  | 9.81E-02 | 1.42 | 2.94-2.16 |  |
|  |  | MR Egger |  | 8.85E-01 | 0.86 | 0.11-6.75 |  |
| HF | ebi-a-GCST009541 | Inverse variance weighted | 36 | 2.32E-04 | 1.42 | 1.18-1.71 | 1.44(1.22-1.70)<br>$p=1.36\text{e-}05$ |
|  |  | Weighted median |  | 1.74E-01 | 1.21 | 0.92-1.59 |  |
|  |  | MR Egger |  | 7.58E-02 | 2.65 | 0.93-7.51 |  |
|  | finn-b-I9_HEARTFAIL | Inverse variance weighted | 40 | 1.92E-02 | 1.52 | 1.07-2.16 |  |
|  |  | Weighted median |  | 1.24E-01 | 1.47 | 0.90-2.41 |  |
|  |  | MR Egger |  | 6.41E-01 | 1.73 | 0.18-17.08 |  |
| MI | ebi-a-GCST011365 | Inverse variance weighted | 36 | 1.77E-06 | 1.60 | 1.32-1.95 | 1.63(1.37-1.94)<br>$p=3.11\text{e-}08$ |
|  |  | Weighted median |  | 6.12E-04 | 1.57 | 1.21-2.02 |  |
|  |  | MR Egger |  | 2.56E-01 | 1.93 | 0.63-5.91 |  |
|  | finn-b-I9_MI | Inverse variance weighted | 40 | 4.81E-03 | 1.75 | 1.19-2.57 |  |
|  |  | Weighted median |  | 3.16E-03 | 2.12 | 1.29-3.50 |  |
|  |  | MR Egger |  | 2.87E-01 | 0.26 | 0.02-3.01 |  |

Notes: Two independent cohorts were employed for primary analysis and we used meta-analysis to evaluate the consistency of these results. In the meta-analysis, when heterogeneity exists ( $p<0.05$ ), a random effect model was used, otherwise, a fixed effect model would be used. Almost all CVDs used fixed effect model to calculate odds ratios, except AF ( $p<0.01$ ). GWAS, Large genome-wide association studies; OR, odds ratio; CI, confidence interval; CAD, coronary artery disease; AF, atrial fibrillation; HF, heart failure; MI, myocardial infarction.

Additionally, we also tried to find any genetically predicted casual effects of different cardiovascular events on mood instability. However, no significant results were shown and these results could be found in **Table S1**.

#### 3.1.2 Robustness of primary analysis

Using the MR-Egger regression and Weighted median method, the results were consistent with IVW method, which identified the reliability of the primary analysis.

There were no horizontal pleiotropic effects in MR-Egger intercept **(Figure 2a-d)** and no single SNP could determine the total causal effects of mood instability on CAD, MI, HF and stroke by using “Leave-one-out” method **(Figure 2e-h)**. MR-PRESSO identified several outlier SNPs in different CVDs respectively **(Table S2)**. After correcting these outliers, significant results were observed in these four CVD events.

**Figure 2.**
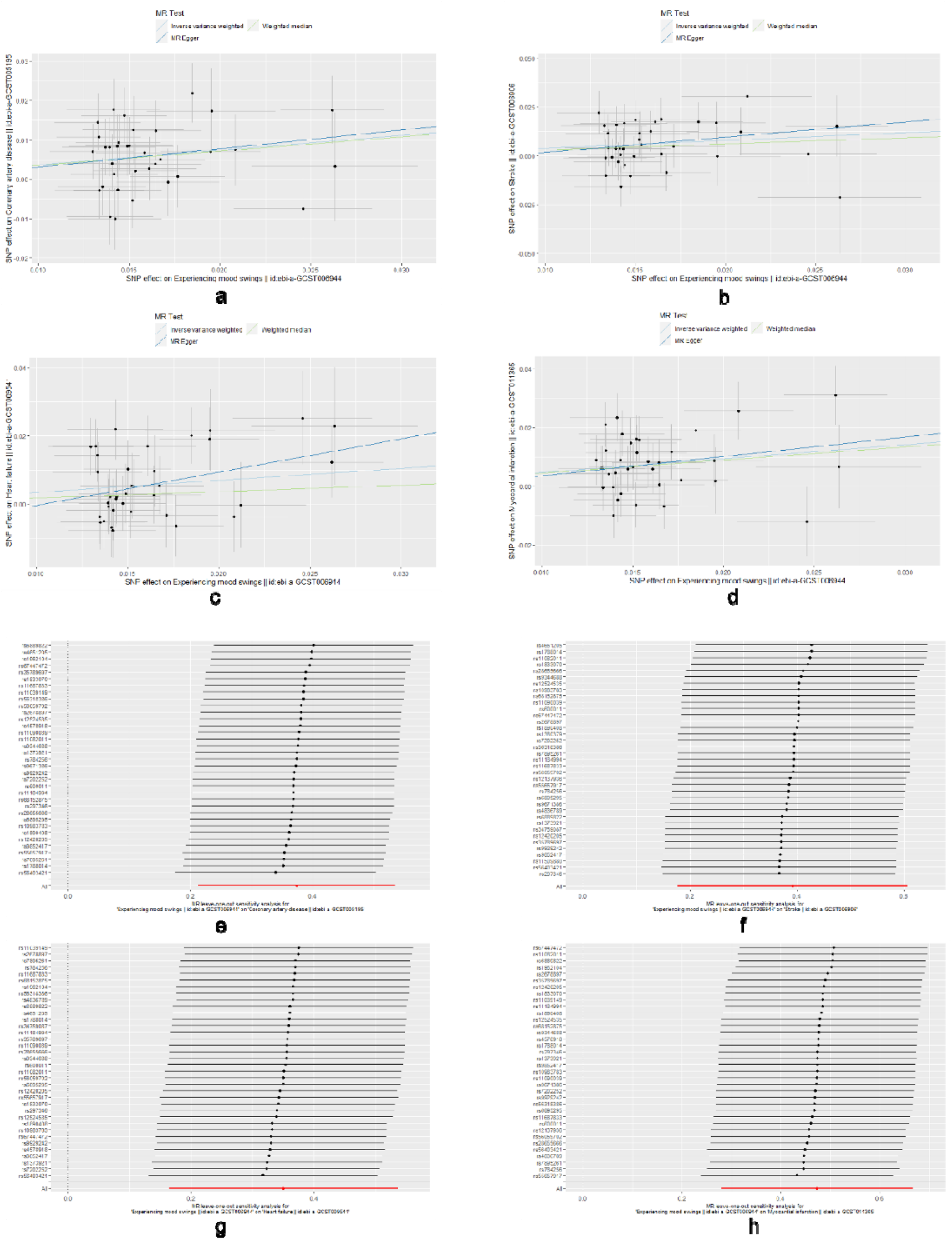
Sensitive analysis about the causal effects of mood instability on different CVDs. Figure 2a-d are scatter plots and each plots represents an IV. The line on each point reflects the 95% CI, and the vertical coordinate is the effect of SNPs on (a) CAD, (b) stroke, (c) HF and (d) MI. The horizontal coordinate is the effect of SNPs on mood swings. The slope of the line corresponds to the causal estimation. A positive slope indicates that mood swings had a positive effect on cardiovascular events. Figure 2e-h are “Leave-one-out” graphs demonstrate that all lines were situated towards the positive side of the “0 line”, suggesting that the removal of any SNPs did not significantly impact the findings including (e) CAD, (f) stroke, (g) HF and (h) MI, implying the robustness of MR results. CVD, cardiovascular disease; IV, instrumental variable; CI, confidence interval; SNP, single nucleotide polymorphism; MI, myocardial infarction; HF, heart failure; CAD, coronary artery disease; MR, mendelian randomization.

### 3.2 MVMR analysis

MVMR was employed for significant outcomes in primary analysis to further investigate the effect of potential confounding factors on causal estimates^33^. In most cases, the causal effects of mood instability on 5 CVDs were remarkably robust **(Table 3)**. However, after adjusting hypertension, the causal estimates were not significant in all circumstances, which suggested that hypertension may play an important role in the pathway from mood instability to CVDs.

**Table 3.** MVMR outcomes

| Potential confounding factors | Outcome | nsnp | Pval | OR | 95%CI |
| --- | --- | --- | --- | --- | --- |
| Hypertension | CAD | 32 | 0.70 | 1.10 | 0.68-1.76 |
|  | MI | 33 | 0.45 | 1.18 | 0.76-1.84 |
|  | HF | 32 | 0.16 | 1.21 | 0.93-1.57 |
|  | Stroke | 33 | 0.27 | 1.19 | 0.87-1.62 |
| BMI | CAD | 27 | 1.52e-03 | 1.47 | 1.16–1.86 |
|  | MI | 27 | 1.57e-03 | 1.51 | 1.17–1.95 |
|  | HF | 27 | 3.92e-02 | 1.33 | 1.01-1.75 |
|  | Stroke | 27 | 1.74e-02 | 1.41 | 1.06-1.88 |
| Drinking | CAD | 32 | 1.55e-03 | 1.53 | 1.18-1.99 |
|  | MI | 33 | 1.51e-03 | 1.59 | 1.19-2.12 |
|  | HF | 32 | 1.54e-02 | 1.37 | 1.06-1.76 |
|  | Stroke | 33 | 5.67e-03 | 1.41 | 1.11-1.81 |
| Smoking | CAD | 38 | 2.89e-04 | 1.49 | 1.20–1.86 |
|  | MI | 39 | 2.69e-04 | 1.63 | 1.25–2.12 |
|  | HF | 38 | 8.58e-03 | 1.34 | 1.08–1.67 |
|  | Stroke | 39 | 2.45e-02 | 1.34 | 1.04–1.73 |

Notes: BMI, body mass index; MI, myocardial infarction; HF, heart failure; CAD, coronary artery disease.

### 3.3 Mediation analysis

Our findings identified that hypertension played a role in the causal effect of mood instability on CAD, MI, HF and stroke. The proportions of mediation were shown in **Table 4**. However, other potential mediators such as IL-6 and CRP have no such mediation effect.

**Table 4.** Mediation analysis outcomes

| Outcomes | Mediators | Proportion (%) | 95%CI |
| --- | --- | --- | --- |
| CAD | Hypertension | 39.60 | 19.31–59.89 |
|  | IL-6 | -0.23 | -10.49–10.02 |
|  | CRP | 0.68 | -1.61–2.96 |
| MI | Hypertension | 35.37 | 17.10–53.65 |
|  | IL-6 | -0.28 | -12.50–11.94 |
|  | CRP | 0.67 | -1.67–3.01 |
| HF | Hypertension | 43.19 | 20.68–65.69 |
|  | IL-6 | -0.17 | -6.35–6.00 |
|  | CRP | 0.12 | -2.23–2.47 |
| Stroke | Hypertension | 55.47 | 27.00–83.95 |
|  | IL-6 | -0.46 | -14.82–13.90 |
|  | CRP | -1.39 | -4.06–1.28 |

Notes: IL-6, Interleukin-6; CRP, C-Reactive protein.

## 4. Discussion

In this MR study, we explored the causal relationships between mood instability and 5 high-frequency CVDs by using large-scale and open GWAS statistics. Three main findings were yield: Firstly, mood instability has a significant causal effect on a wide range of CVDs including CAD, MI, HF and stroke, which has been identified robust in sensitive analysis. However, no evidence claimed that mood instability could increase the risk of AF. Secondly, in most cases, common risk factors of CVD could not affect this causal effect, emphasizing the profound influence caused by mood instability. Thirdly, complementary analysis indicated hypertension may play an important role in the causal pathway of mood instability to CAD, MI, HF and stroke. Finally, we did not observe any statistically significant results in the reverse MR analysis.

Personality traits, especially mood instability (mood swings), which start to have an enduring impact from birth, the causal relationship to cardiovascular events are rarely investigated. Many researches have found MDD, anxiety disorder and bipolar disorder may accelerate the progression of CVD. A meta-analysis including 26 studies of 1,957,621 individuals revealed that depression increased the risk of stroke (HR 1.13; 95% CI 1.00-1.28), MI (HR 1.28; 95% CI, 1.14-1.45), HF (HR 1.04; 95% CI, 1.00-1.09) as well as the risk of cardiovascular morality (HR 1.44; 95% CI, 1.27-1.63)^6^. In the China Kadoorie Biobank (CKB) cohort study involving 512,712 persons, depression significantly increased CVD mortality (HR, 1.39 95%CI, 1.10-1.76), especially in male^8^. Previously two-sample mendelian randomization analysis conducted by Lei Wang^9^ et al. found the potential causal relationship between depression and AF (OR = 1.098, 95% CI 1.000–1.206; P = 0.049). Another MR study found mood disorders (MDD and BD) causally resulted in both stroke (OR 1.07; 95% CI 1.03–1.11) and ischemic stroke (OR 1.09; 95% CI 1.04–1.13)^16^. A cohort study conducted by Shinya Nakada et al. demonstrated that the risk of CVD among patients with anxiety disorder was higher than those without mental illness (HR 1.72; 95% CI 1.32–2.24)^7^. Similarly, the prevalence of CVD is significantly greater among BD patients than controls (OR 4.95; 95% CI 4.27–5.75)^34^. Several MR studies focused on the causal association between neuroticism and CVD draw negative results. Gull Rukh et al.^35^ found neuroticism could not increase the risk of any CVD traits (p>0.05). Among diabetes mellitus individuals, neuroticism also has no causal effects on CAD^15^. To best of our knowledge, this is the first study using MR to investigate the causal association between mood instability and different cardiovascular endpoints. Furthermore, these significant results clarified not only mental illness, but also personality traits could render a considerable powerful impact on circulatory system.

However, the pathophysiological mechanisms connecting mood swings to the progression of CVD is still unclear. Here, we proposed three potential mechanisms about how mood instability contributes to the development of CVDs. Firstly, psychiatric traits are associated with oxidative stress^36^. During the process of oxidative stress, total bilirubin (TBil) plays a vital role in antioxidant activity and cellular protection^37^. Mood instability may decrease TBil level and the lack of TBil brings to the damage to vascular endothelial cells, leads to hypertension in the end^38^. Persistent hypertension causes artery atherosclerosis and myocardial remodeling, which increase the risk of cardiovascular events. Interestingly, mood instability can cause hypertension, which is consistent with the result in our mediation analysis. Secondly, mood instability causes dysregulation of endocrine function. Glucocorticoids can reduce inflammation under a normal condition. However, long-term emotional instability can lead to abnormal secretion of glucocorticoids. Under the inhibition of high-concentration glucocorticoids, the immune system becomes insensitive, leading to an increased susceptibility to CVDs^39^. Thirdly, mood instability can lead to mental stress^40^, and studies have identified that cardiac sympathetic nerves can be abnormally activated during long-term mental stress, which can decrease the blood flow, increase heart rate and cause ventricular hypertrophy, leading to CVDs^6,41^.

Given that mood instability, this personality trait is an upstream factor to mental illness and have significant causal effects on CVDs, a focus on early recognition and management of mood swings may potentially lower the incidence of both psychotic disorder and cardiovascular disease. Although there is no direct evidence identified this hypothesis, some randomized controlled trails (RCT) aiming at depression and CVDs have draw some conclusion. They found that using selective serotonin reuptake inhibitors (SSRIs) in CAD patients with depressive symptoms was safe and effective ^42,43^. However, further researches are needed to clarify whether an early intervention to mood instability can change cardiovascular outcomes. Additionally, as mood instability is a very common personality trait among normal crowds, it may be necessary for clinicians to evaluate mental health at regular intervals by using questionnaire such as Mood Zoom (MZ),which has been proved a standard measurement for depression, anxiety and mental health^44^. Furthermore, a recent RCT demonstrated that stress related disorders were strongly associated with early onset CVD^45^, and regular exercise seems very important for releasing pressure as well as lowering the risk of CVD^30,31^. So besides regular screening, encouraging people with mood instability to exercise is also meaningful. Moreover, Niall M. McGowan^46^ further explored the relationship between mood instability and blood pressure (BP), they found BP may be a valuable predictor of mood instability, which emphasized the importance of Home blood pressure monitoring (HBPM) at regular intervals. The management of BP is likely to become a potential explicit treatment target for both improving unhealthy emotion and preventing CVDs.

Still, further studies remain to be conducted to develop a more comprehensive model for the management of physical and mental illnesses.

Our novel MR study had several strengths. As mentioned above, this study is the first to comprehensively estimate the casual relationship between mood instability (mood swings) and a wide range of cardiovascular endpoints. Additionally, for CVDs, two representative and independent cohorts were employed to ensure the causal results stable. Furthermore, most of SNPs have passed F-test (F>10), which showed a strength to be IVs for exposure. Then, we used various of methods for sensitive analysis and identified the robustness of our outcomes. Moreover, in order to eliminate common confounding factors which may affect the direction of causal results, we conducted complementary analysis by means of MVMR and mediation MR. Ultimately, we found hypertension mediated the causal effects from mood instability to CVDs, so we further investigated relevant mechanisms for clinical reference.

Some limitations can not be ignored. Firstly, statistics utilized in this study was from European ancestry, which constrained further extrapolation of the results to other racial groups. Secondly, we only selected one personality trait in this MR study, and other traits need to be included in the further studies. Thirdly, there may still be some unrecognized pathways between the exposure and outcome variables, potentially introducing biases into our results.

## 5. Conclusion

In conclusion, this bidirectional two-sample MR analysis is the earliest study to identify that mood instability has significant causal effects on CVDs including CAD, MI, HF and stroke, which can be partly mediated by hypertension. Enhancing emotional management and controlling blood pressure may be beneficial for decreasing the risk of CVDs, which need more support of evidence-based medicine.

## Data Availability

All data produced in the present study are available upon reasonable request to the authors
All data produced in the present work are contained in the manuscript
All data produced are available online at https://gwasmrcieu.ac.uk/

## 6. Declarations

### 6.1 Ethics approval and consent to participate

Not applicable.

### 6.2 Consent for publication

Not applicable.

### 6.3 Availability of data and materials

All data generated or analysed during this study are included in this article and its supplementary information files.

### 6.4 Competing interests

The authors declare that they have no competing interests.

## 7. Fundings

The study was funded by the Suzhou Science and Technology Project (SKYD2022103), Suzhou Specialized Program for Diagnosis and Treatment Techniques of Clinical Key Diseases (LCZX202103), and Soochow University (P112206422).

## 8. Supplementary materials

Additional file 1, Table S1, Causal associations of CVDs on mood instability; Figure S1. Study design and 3 assumptions of MR analysis. Table S2. MR-PRESSO global test of primary analysis.

## Notes

### Competing Interest Statement

The authors have declared no competing interest.

### Author Declarations

All the summary statistics were publicly available which obtained from the Open GWAS database developed by the MRC Integrative Epidemiology Unit (IEU) (https://gwasmrcieu.ac.uk/).

