## Supplementary material for "Mood instability may be causally associated with the high risk of cardiovascular disease: evidence from a mendelian randomization analysis": Figure S1

***Supplementary materials***

Zirui Liu, Haocheng Wang, Zhengkai Yang, Yu Lu, Cao Zou

**Contents**

**Figure S1.** Study design and 3 assumptions of MR analysis.

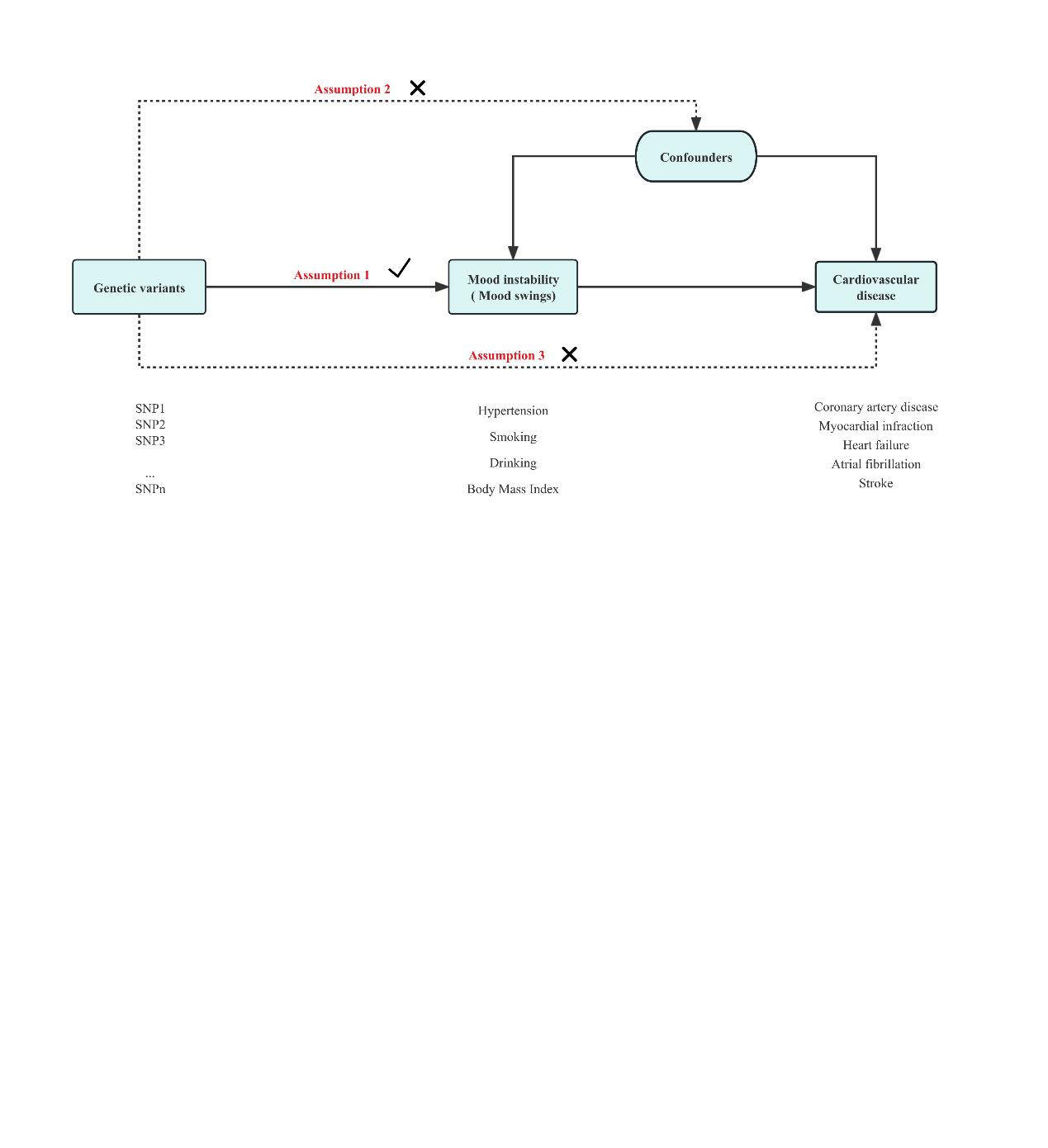

Figure S1. Study design and 3 assumptions of MR analysis.
