## Supplementary material for "Mood instability may be causally associated with the high risk of cardiovascular disease: evidence from a mendelian randomization analysis": Table S1

***Supplementary materials***

Zirui Liu, Haocheng Wang, Zhengkai Yang, Yu Lu, Cao Zou

**Contents**

**Table S1.** Causal associations of CVDs on mood instability.

Table S1. Causal associations of CVDs on mood instability.

| **Exposure** | **Method** | **nsnp** | **Pval** | **OR** | **95%CI** |
| --- | --- | --- | --- | --- | --- |
| CAD  ebi-a-GCST005195 | Inverse variance weighted | 61 | 0.35 | 1.01 | 0.99-1.02 |
|  | Weighted median |  | 0.31 | 1.01 | 0.99-1.03 |
|  | MR Egger |  | 0.17 | 0.98 | 0.95-1.01 |
| AF  ebi-a-GCST006414 | Inverse variance weighted | 102 | 0.92 | 1.00 | 0.99-1.01 |
|  | Weighted median |  | 0.58 | 1.00 | 0.99-1.02 |
|  | MR Egger |  | 0.86 | 1.00 | 0.98-1.02 |
| Stroke  ebi-a-GCST006906 | Inverse variance weighted | 8 | 0.42 | 0.98 | 0.93-1.03 |
|  | Weighted median |  | 0.84 | 1.00 | 0.95-1.04 |
|  | MR Egger |  | 0.73 | 1.08 | 0.73-1.59 |
| HF  ebi-a-GCST009541 | Inverse variance weighted | 8 | 0.95 | 1.00 | 0.95-1.05 |
|  | Weighted median |  | 0.98 | 1.00 | 0.96-1.04 |
|  | MR Egger |  | 0.46 | 0.95 | 0.83-1.08 |
| MI  ebi-a-GCST011365 | Inverse variance weighted | 77 | 0.15 | 1.01 | 1.00-1.02 |
|  | Weighted median |  | 0.06 | 1.01 | 1.00-1.03 |
|  | MR Egger |  | 0.47 | 0.99 | 0.96-1.02 |

Notes: OR, odds ratio; CI, confidence interval; CAD, coronary artery disease; AF, atrial fibrillation; HF, heart failure; MI, myocardial infarction.
