## Supplementary material for "Mood instability may be causally associated with the high risk of cardiovascular disease: evidence from a mendelian randomization analysis": Table S2

***Supplementary materials***

Zirui Liu, Haocheng Wang, Zhengkai Yang, Yu Lu, Cao Zou

**Contents**

**Table S2.** MR-PRESSO global test of primary analysis.

Table S2. MR-PRESSO global test of primary analysis.

| **Phenotypes** | **global *P*** | **Outliers SNP** | **global *P* after removing outliers** |
| --- | --- | --- | --- |
| CAD | <0.001 | rs11509880, rs1360379, rs34759087, rs4836789 | 0.51 |
| MI | <0.001 | rs11509880, rs1360379, rs4651205 | 0.16 |
| HF | 0.032 | rs11509880, rs1360379, rs9671386 | 0.42 |
| Stroke | 0.018 | rs11039149, rs1962104, rs4578918 | 0.56 |
